# Towards Interpretable AI Second Opinions: Foundation Model Heatmaps in Radiology

**DOI:** 10.64898/2026.08.20.26360908

**Authors:** Ethan Dack, Chengliang Dai, Hanno Hoppe, Paula Krüselmann, Stefanie Meiler, Warissara Jutidamrongphan, Ling Wang, Kun Tang

## Abstract

AI-assisted diagnostic tools typically act as a “second opinion,” providing radiologists with a discrete prediction or probability score that can be consulted alongside clinical context. This treats AI as an independent advisor rather than a collaborative partner, leaving its reasoning largely opaque. We explore a complementary approach grounded in human-AI collaboration through visual interpretability. Specifically, we investigate (1) radiologist performance when diagnosing chest X-rays from images alone, and (2) whether deep learning-generated heatmaps can support radiologists *during* this diagnostic process, rather than merely validating a final answer. We developed an interactive application that enables readers to engage directly with model-generated heatmaps as they form their diagnoses, and conducted a user study to evaluate how this influences diagnostic behaviour and accuracy. Our findings offer new insights into integrating interpretable, spatially grounded AI feedback into radiologist workflows. Code, datasets, and the application can be found at https://github.com/eedack01/heatmap_assisted_diagnosis/.

## 1 Introduction

Radiologists routinely use chest X-ray (CXR) imaging for diagnosis, analysing images alongside available clinical information [3]. To increase diagnostic confidence, radiologists sometimes seek a second opinion [12]. Recently, artificial intelligence (AI) systems have increasingly been positioned as this second opinion [35]. In this role, AI often provides a binary classification of disease presence. While this can enhance operational efficiency and reduce costs, it risks fostering automation bias among radiologists. If clinicians become overly reliant on these outputs, diagnostic vigilance may decrease, increasing the likelihood that subtle or complex findings are overlooked [2,27].

Driven by self-supervised methods [14,8,7], foundation models have become central to computer vision [5]. They are particularly important in radiology, where Vision Transformers (ViTs) [11] are difficult to train from scratch [32] and labelled data are often limited [25]. These models are expected to learn robust, transferable representations [25,5], yet what these representations capture remains difficult to interpret. Heatmaps offer one route to this interpretability. A visualisation technique used for over a century [37], and popularised in deep learning by gradient-based methods such as Grad-CAM [30,39] for highlighting image regions most relevant to a model’s prediction. We apply Grad-CAM to fine-tuned foundation models to interpret their learned representations.

While such heatmaps are typically used to interpret model behaviour, visual representations have also been shown to improve disease classification performance in AI models themselves [1,31]. This raises a natural question: *can these same visual explanations support human diagnostic performance?* [28]. Highlighting the regions contributing to a foundation model’s prediction offers radiologists additional insight, not only for disease classification but also for educational purposes [22,29]. Since foundation models are trained on large-scale image datasets, their learned representations may, in theory, encode visual information useful for supporting radiologists during image interpretation.

By integrating heatmap outputs from foundation models into radiology workflows, this paper investigates whether foundation model-generated heatmaps can assist radiologists and improve diagnostic performance. To this end, we designed a human–AI collaboration study [33,6] and developed an application that enables radiologists to interpret CXR images with the assistance of heatmaps derived from foundation models. The study workflow is depicted in Figure 1. First, a radiologist makes an initial diagnosis. Second, the same radiologist reviews the image alongside the corresponding heatmaps and provides a second diagnosis. Using this setup, we collected diagnostic predictions from five radiologists with varying levels of experience, resulting in a curated dataset that we release for research purposes.

**Fig. 1.**
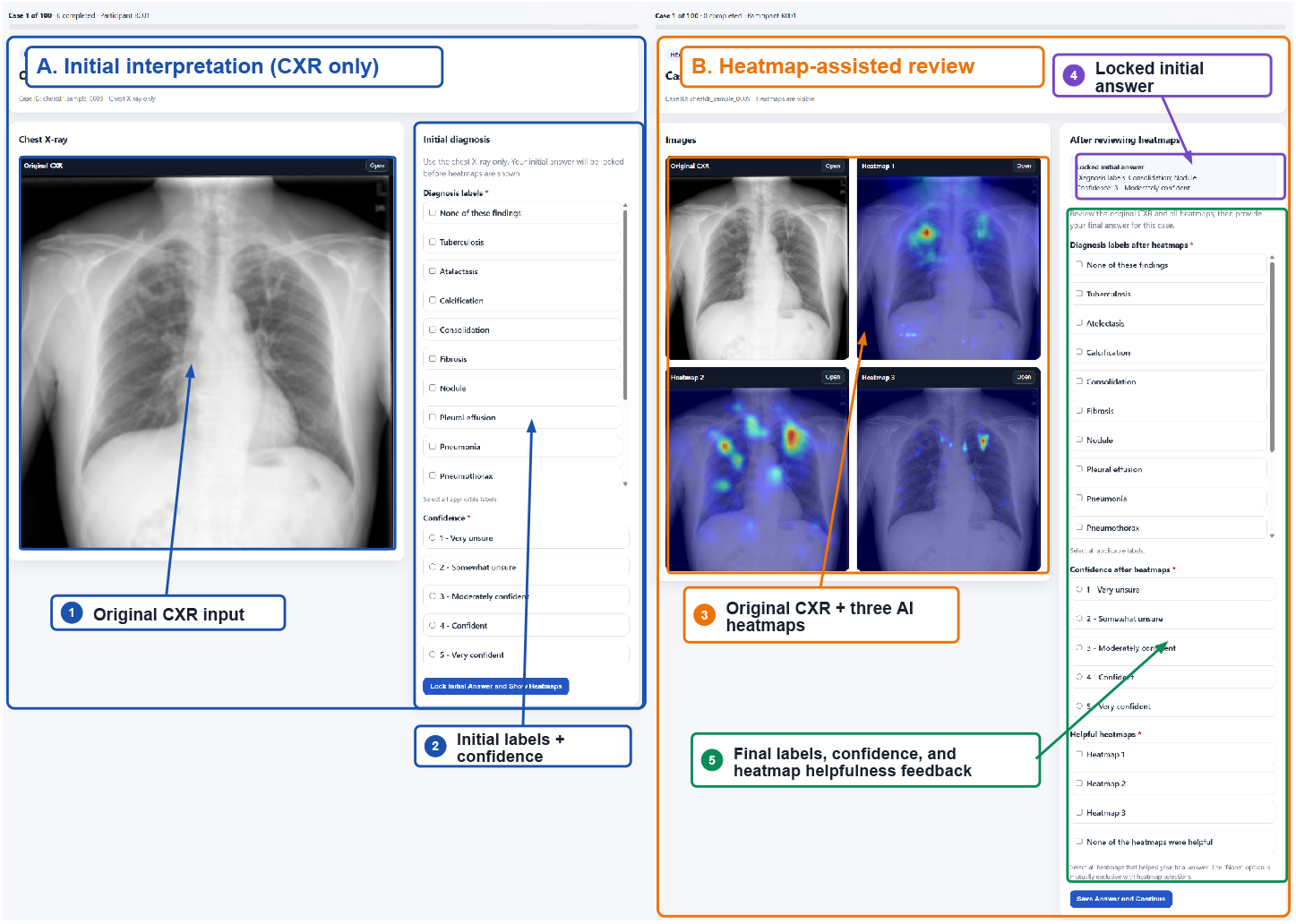
Screenshot of the interactive tool. Study workflow – Initial response is locked before heatmaps. Participants then review the heatmaps, submit a final diagnosis, and indicate which heatmaps were helpful.

## 2 Datasets and Models

### 2.1 Datasets

We selected two datasets, VinDr and ChestDr [23,36], with clinically relevant diagnostic tasks and board-consensus test-set labels. The tasks are challenging for both human readers and AI models due to the large number of classes and class imbalance. We selected 50 samples from each dataset. In consultation with radiologists, we identified a subset of labels of high clinical interest and ensured that at least one of these labels was represented among the 50 samples selected from each dataset. This sampling strategy avoids representing classes, such as cardiomegaly, that appear easier to diagnose for both humans and AI models [4,19]. Both datasets rely solely on human annotations, unlike approaches that use NLP-based label extraction [16,17].

### 2.2 Models

We selected recent foundation models that have demonstrated promising performance. All three selected foundation models are based on similar ViT architectures and contain a comparable number of parameters. We focus on the version of **ARK** [21] trained with an image size of 224 *×* 224; it is pretrained using a teacher-student cyclical framework and initialised with ImageNet [10] weights to extract knowledge from the available datasets. **EVA-X** [38] combines contrastive learning and masked image modelling [14] during pretraining. **RAD-DINO** [26] is directly inspired by DINOv2. It initialises the model with DINOv2 [24] weights and applies domain transfer.

## 3 Study Design

### 3.1 Foundation Model Fine-tuning and Heatmap Generation

We opt for full fine-tuning rather than linear probing, as this yielded improved performance and more interpretable heatmaps. Each foundation model is fine-tuned separately for each task within each dataset, resulting in six trained and evaluated models. Foundation models can be susceptible to catastrophic forgetting [18]; therefore, we perform a small hyperparameter search over the learning rate for each model, selecting from 1 *×* 10^−5^, 5 *×* 10^−5^, 1 *×* 10^−4^. Inspired by the original Grad-CAM papers [39,30], we generated heatmaps using a per-token “gradient *×* activation” saliency map, adapted from CNN feature maps to ViT patch tokens, with averaging performed across both layers and predicted classes. The best-performing model was selected based on validation performance and qualitative inspection of the generated heatmaps.

## 4 Results

### 4.1 Interactive Radiology Tool

We recruited five radiologists with varied backgrounds, all of whom had experience with chest CXRs. Table 1 summarises the differences between these experts.

**Table 1.** Radiologist reader characteristics.

| Reader | Training | Experience | CXR/year |
| --- | --- | --- | --- |
| R001 | Radiology resident | 2 years | ~18,000 |
| R002 | Cardiovascular Fellowship | 8 years | ~5,000 |
| R003 | Cardiovascular Fellowship | 27 years | ~10,000 |
| R004 | General radiology | 14 years | ~20,000 |
| R005 | General radiology | 7 years | ~10,000 |

For this study, we implemented an interactive tool using AI-assisted coding practices [13] to track radiologist results and generate the dataset. Figure 1 illustrates the developed application, highlighting the key components of the user interaction. The application presents two screens per CXR, one for recording the initial diagnosis and one for recording the diagnosis after heatmap review, along with confidence levels for both, and allows radiologists to flag heatmaps they found useful. These annotations are logged for each user, enabling analysis of intra-rater variability.

We conducted a paired-reader study to evaluate whether Grad-CAM heatmaps influence radiologists’ diagnostic decisions for CXRs. The radiologists independently reviewed 100 chest radiographs. For each case, the radiologist first provided a multi-label diagnosis based solely on the radiograph. The same case was then shown again with three Grad-CAM heatmaps generated by the foundation models, and the radiologist provided a second diagnosis. This produced 500 paired reader-case observations before and after the heatmap presentation.

The study used a within-reader, within-case design: each radiologist served as their own control, with paired CXR-only and heatmap-assisted ratings for every case. Our main research question was whether heatmap access changed diagnostic accuracy, overlap with reference labels, confidence, or selected diagnosis categories. The task was intentionally difficult, requiring radiologists to infer diagnoses from the image alone, without clinical history, blood tests, microbiology, or other context, so strict exact agreement with the full reference label set was expected to be low. To keep the analysis clinically interpretable, we used both strict original-label metrics and pooled grouped-label metrics.

The foundation models have been extensively tested in their respective papers on open-source datasets [38,21,26]. We evaluated them here using macro-F1, Hamming loss, AUROC, and AUPRC, retaining both the best-performing and last training checkpoints. Mean AUROC values for ChestDr and VinDr, respectively, were 0.821 and 0.885 for **ARK**, 0.818 and 0.867 for **EVA-X**, and 0.839 and 0.868 for **RAD-DINO**. Since predictive performance does not necessarily yield interpretable or reliable heatmaps [15,20], we assessed the hyperparameter search by generating five heatmaps per model and dataset (180 total), visually inspecting them for spurious correlations, and selecting models with the fewest observed spurious cases.

Given the difficulty of the task, we analyse the results twice: once with strict labels and once with pooled labels. The primary pooled analysis used the following predefined diagnosis groups:

– **Airspace/infection**: Pneumonia, Consolidation, Lung opacity, Atelectasis
– **Pleural abnormality**: Pleural effusion, Thickened pleura
– **Chronic TB/fibrotic/cavity**: Tuberculosis, Fibrosis, Lung cavity
– **Nodule/calcified lesion**: Nodule, Calcification
– **Pneumothorax**
– **Pulmonary edema**

The label “None of these findings” was treated as mutually exclusive. If it co-occurred with any abnormal finding in a response, it was removed prior to metric calculation. Paired changes in grouped Jaccard and confidence were assessed with Wilcoxon signed-rank tests and case-cluster bootstrap confidence intervals; selection-shift tests used Benjamini-Hochberg FDR correction.

Figure 2 shows the primary endpoint, grouped Jaccard overlap, between the radiologist-selected and reference diagnosis groups. This metric was chosen because it gives partial credit in this difficult multi-label task. Grouped and strict exact-match accuracies were reported only as descriptive measures of task difficulty. Across 500 reader-case pairs, grouped Jaccard increased slightly from 0.404 to 0.418 (mean difference 0.014; Wilcoxon p = 0.042; case-cluster bootstrap 95% CI -0.0004 to 0.0291). We observe a general pattern in radiologists’ diagnostic performance across difficulty levels. In certain cases, the heatmap helped confirm the correct diagnosis.

**Fig. 2.**
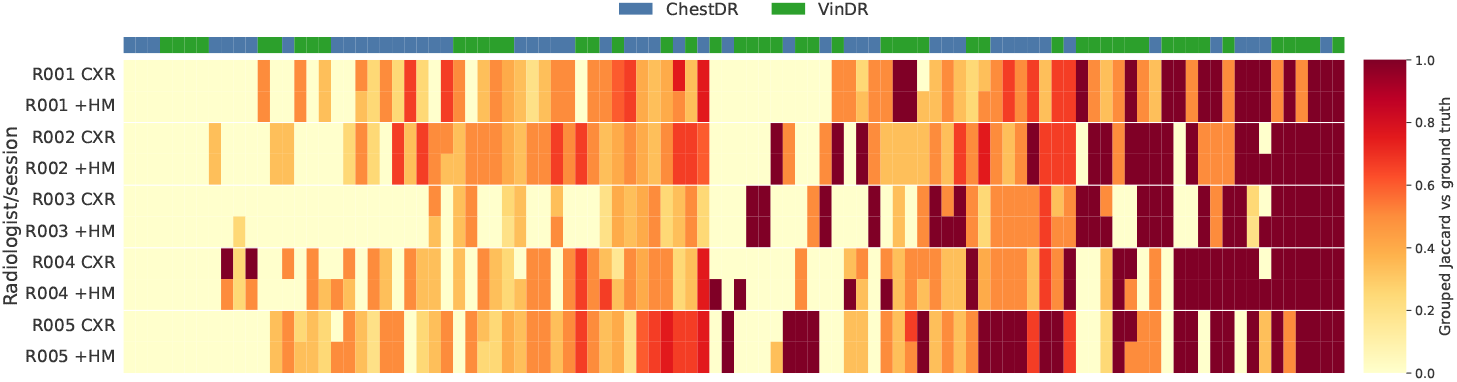
Grouped Jaccard by case before and after heatmaps. Rows show each radiologist and session; **+HM** denotes the second diagnosis with heatmap assistance. Cases are sorted by grouped difficulty, and strong red indicates an exact match.

As demonstrated in Figure 3, we also report per-group/per-label F1 scores and selection shifts, defined as the number of reader-case pairs in which a diagnosis group, label, or response option was added after heatmap presentation minus the number in which it was removed. Exact-match accuracy remained low and was used only as context: grouped exact accuracy changed from 19.2% to 20.2%, and strict diagnostic-label exact accuracy from 7.2% to 7.8%. Heatmaps changed the grouped diagnosis set in 9.8% of reader-case pairs and the original response set in 13.0%. Per-group F1 changes were small, and inter-reader agreement did not increase after heatmap presentation. The most consistent behavioural shift was away from the response-only “None of these findings” option (net -16; FDR-adjusted p = 0.00058) and toward abnormal findings, particularly Airspace/infection (net +14; FDR-adjusted p = 0.00058) and Nodule/calcified lesion (net +11; FDR-adjusted p = 0.034).

**Fig. 3.**
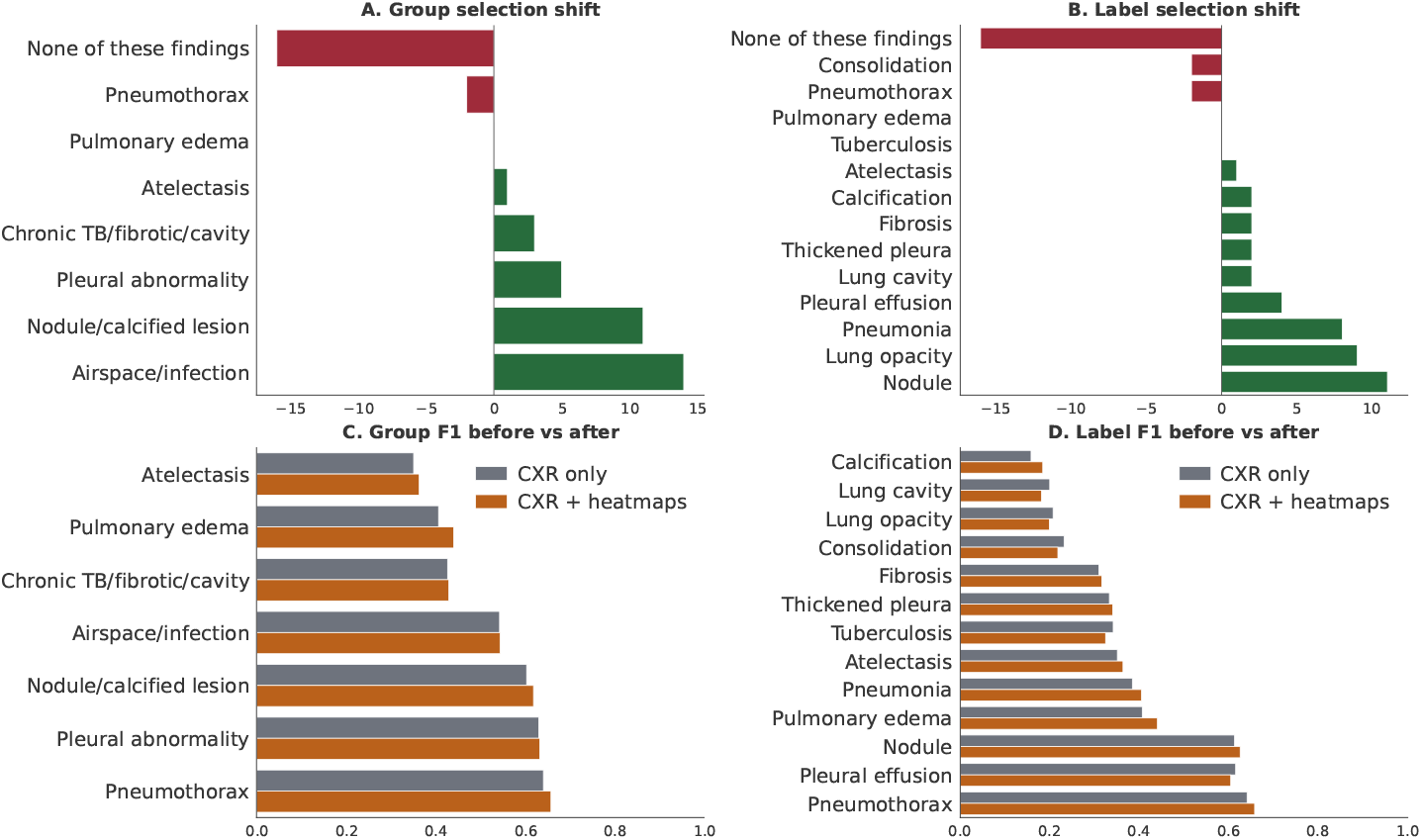
F1 scores and selection shift. Panels (a–b) show selection shifts, instances where a radiologist added or changed a diagnosis following heatmap presentation. Panels (c–d) show F1 scores before and after heatmap presentation.

Radiologist confidence is shown in Figure 4: mean confidence increased from 3.64 to 3.70 (mean difference 0.06; Wilcoxon p = 0.0034; bootstrap 95% CI 0.012 to 0.106). Table 2 shows that **EVA-X** was rated most helpful and was associated with the largest confidence increase, while **RAD-DINO** produced the largest gains in diagnostic performance. Table 3 reveals substantial variance in individual radiologist preferences: reader 005 found only 29 useful heatmaps, compared to 110 for reader 003. This variation in usefulness across models likely stems from the regions each model highlighted: **RAD-DINO** produced very small highlighted areas, **ARK** highlighted much larger areas, and **EVA-X** fell in between.

**Fig. 4.**
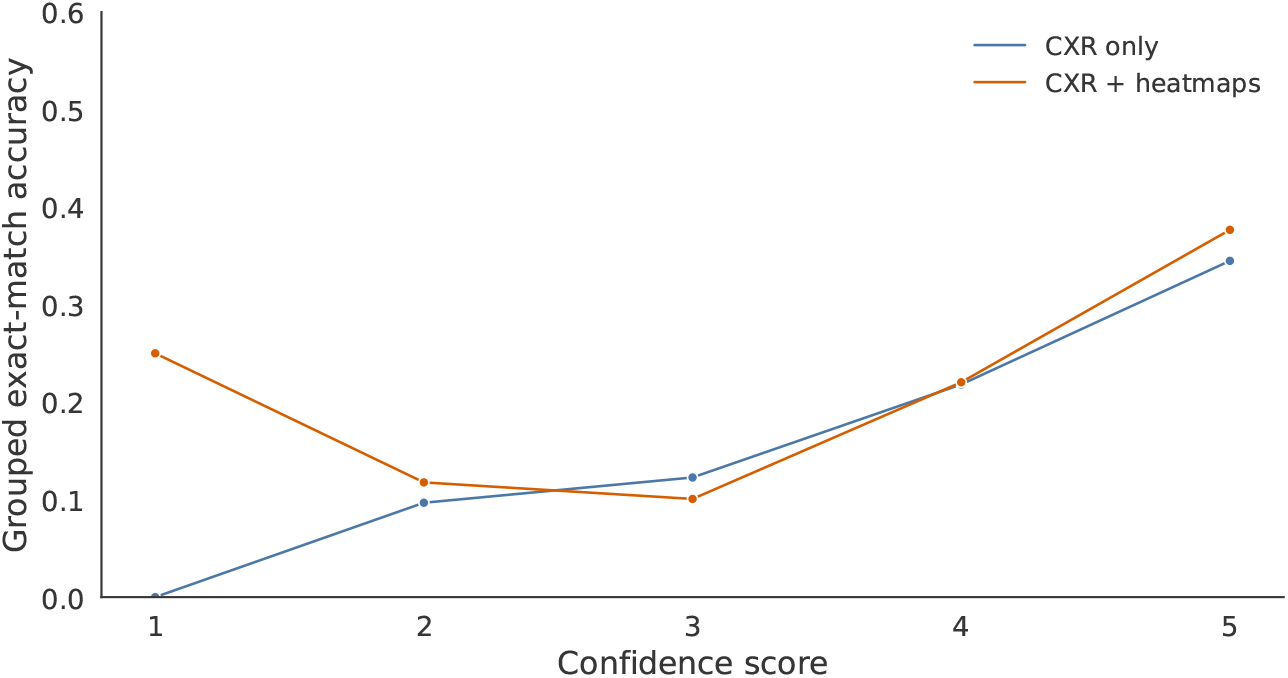
Grouped accuracy by confidence level. Heatmaps improved accuracy more in cases for which radiologists were less confident.

**Table 2.** Model-level summary of selected reader–case reviews.

| Model | Selected reviews | Reviews (%) | Mean grouped Jaccard $\Delta$ | Mean conf. $\Delta$ | Conf. increased |
| --- | --- | --- | --- | --- | --- |
| **EVA-X** [38] | **128** | **25.6** | +0.013 | **+0.266** | 34/128, 26.6% |
| **RAD-DINO** [26] | 112 | 22.4 | **+0.064** | +0.170 | 25/112, 22.3% |
| **ARK** [21] | 78 | 15.6 | +0.014 | +0.179 | 15/78, 19.2% |

**Table 3.** Radiologist–Foundation Model Heatmap Interaction Summary.

| Radiologist | Helpful reader-case reviews | Total useful heatmaps | ARK | EVA | RAD-DINO |
| --- | --- | --- | --- | --- | --- |
| R001 | 29 | 36 | 7 | 17 | 12 |
| R002 | 53 | 67 | 17 | 26 | 24 |
| R003 | 62 | 110 | 34 | 47 | 29 |
| R004 | 65 | 76 | 13 | 28 | 35 |
| R005 | 26 | 29 | 7 | 10 | 12 |

## 5 Discussion

AI is still far from replacing radiologists; despite groundbreaking progress, it continues to struggle with fine-grained tasks [7]. In this difficult multi-label CXR diagnosis task, Grad-CAM heatmaps yielded only a small increase in partial diagnostic overlap with reference labels, without improving overall diagnostic accuracy. Grouped Jaccard, which gives partial credit for identifying the correct diagnostic family, showed a small improvement in paired scores, though the case-cluster bootstrap interval narrowly included zero, so this finding is suggestive rather than definitive.

The main behavioural effect of heatmaps appears to be a shift in radiologists’ decisions rather than a large increase in exact diagnostic accuracy. We did not inform radiologists that the “None of these findings” label was not present within the dataset. Heatmaps nevertheless substantially reduced selection of the “None of these findings” option and increased selection of abnormal findings. This indicates that heatmaps encouraged radiologists to attend to suspicious image regions and select more active pathology labels. Confidence increased after the heatmap presentation, despite only modest changes in accuracy. This is important because AI explanations can influence reader certainty even when objective correctness changes only slightly. The lack of increased inter-reader agreement further suggests that heatmaps did not produce uniform convergence in decisions; their effect was reader- and case-dependent.

This study has certain expected limitations. First, since heatmaps are class-dependent, incorrect model predictions can yield false-positive heatmaps to radiologists, though this may also prompt them to question their own decisions rather than blindly agree with model outputs. Second, CXRs provide only two-dimensional preliminary assessments: the heart and spine can obscure lesions without a 3D volume, and certain classes, such as lung opacity, are easily confused with others. We observed that fine-tuning foundation models could overcome biases inherited from their training data [9], which may have contributed to better results than training from scratch or using other foundation models with different patch sizes [34,32].

## 6 Conclusion

Overall, the current evidence supports a cautious conclusion: Grad-CAM heatmaps had a measurable but modest impact on radiologists’ diagnostic decisions. They were associated with slightly greater diagnostic overlap within groups and higher confidence, and they shifted selections away from the “None of these findings” response option toward abnormal findings. However, they did not substantially improve exact multi-label diagnostic accuracy.

## Data Availability

Radiologist results are available and a link is posted in the paper.
Heatmaps are also available at the same link posted in the paper.
Application developed is also available at the link posted in the paper.

https://github.com/eedack01/heatmap_assisted_diagnosis

## Acknowledgments

This research was conducted at the University of Bern and UCB. We thank all radiologists who participated in this study. We express our gratitude to Shelley Zixin Shu and Haozhe Luo for insightful discussions throughout this study. Calculations were performed on UBELIX (https://www.id.unibe.ch/hpc), the HPC cluster at the University of Bern.

## Disclosure of Interests

Chengliang Dai is an employee of UCB and may hold shares and/or stock options in UCB.

## Notes

### Competing Interest Statement

The authors have declared no competing interest.

### Author Declarations

VinDr chest X-ray dataset available at: https://physionet.org/content/vindr-cxr/1.0.0/annotations/#files-panel ChestDr chest X-ray dataset available at: https://springernature.figshare.com/articles/dataset/ChestDR_Thoracic_Diseases_Screening_in_Chest_Radiography/22302775?file=39673366

